# Decision support system to evaluate VENTilation in the Acute Respiratory Distress Syndrome

**DOI:** 10.1101/2023.09.28.23295668

**Authors:** Brijesh Patel, Sharon Mumby, Nicholas Johnson, Emanuela Falaschetti, Rhodri Handslip, Sunil Patel, Teresa Lee, Martin S Andersen, Ian M Adcock, Danny McAuley, Masao Takata, Thomas Staudinger, Dan S. Karbing, Matthieu Jabaudon, Peter Schellongowski, Stephen E. Rees, the DeVENT study group

## Abstract

**Rationale:** The acute respiratory distress syndrome (ARDS) shows significant heterogeneity in responsiveness to changes in mechanical ventilation and lacks personalisation.

**Objectives:** Investigate the clinical efficacy of a physiologic-based ventilatory decision support system (DSS) on ARDS patients.

**Methods:** An international, multi-centre, randomized, open-label study enrolling patients with ARDS during the COVID-19 pandemic. The primary outcome was to detect a reduction in average driving pressure between groups. Secondary outcomes included several clinically relevant measures of respiratory physiology, ventilator free days; time from control mode to support mode; number of changes in ventilator settings per day; percentage of time in control and support mode ventilation; ventilation related and device related adverse events; and number of times the advice is followed.

Measurements and Main Results. 95 patients were randomized to this study. The DSS showed was no effect in the average driving pressure between arms. Patients in the intervention arm had statistically improved oxygenation index when in support mode ventilation (−1.41, 95% CI: −2.76, −0.08; p=0.0370). Ventilatory ratio was also significantly improved in the intervention arm for patients in control mode ventilation (−0.63, 95% CI: −1.08, −0.17, p= 0.0068). The application of the DSS resulted in a significantly increased number of ventilator changes for pressure settings and respiratory frequency.

**Conclusions:** The application of a physiological model-based decision support system for advice on mechanical ventilation in patients with COVID-19 and non-COVID-19 ARDS showed that application of about 60% of advice improved physiological state, despite no significant difference in driving pressure as a primary outcome measure.

## Introduction

The clinical presentation of the acute respiratory distress syndrome (ARDS) is highly heterogenous, with varying degrees and types of abnormalities in pulmonary gas exchange and mechanics, and heterogeneity of treatment response to ventilator changes (1, 2). This physiological heterogeneity is highlighted even within a single etiology such as coronavirus disease 2019 (COVID-19) making it increasingly important that ventilator management be tailored to the individual patient (3–8). Ventilator interventions should, however, also be consistent, in so much that physiological phenotypes having similar treatment responses be treated homogenously (9). A strategy of individual but replicable ventilatory care requires understanding of the patient’s physiology at any given moment and standardization of response to individual physiological situations. Currently it has been shown this is not always the case, with 69% of ventilator settings being shown to be non-adherent to current evidence-based lung protective ventilation strategies (10).

Decision support systems (DSS) can aid in such processes of care. When such systems reflect the underlying individual physiology of the patient and provide advice based on this, then care can be seen as being both individualized and replicable (9). The need for such systems to evaluate the current state of physiology is particularly important in complex pathologies such as ARDS. The Beacon Caresystem (Mermaid Care A/S, Nørresundby) is a physiologic based DSS for suggesting changes in mechanical ventilation (11). The system integrates measurements of pulmonary mechanics, gas exchange, indirect calorimetry, volumetric capnography, blood acid-base status, and pulse oximetry to tune mathematical models of patent physiology and provides advice to minimize the risks of negative outcomes. The physiological models of the system have been validated previously including in ARDS patients with and without COVID-19, and the system has been shown to improve physiological state in short periods of mechanical ventilation and reduce pressure support with overly stressing respiratory muscles (12, 13).

This multi-centre study investigates application of a DSS in an ARDS population with varying severities to determine whether application of a DSS could change the physiological status for ARDS patients, and evaluate the barriers to implementation.

## Methods

### Trial Design and Oversight

DeVENT was a multi-centre, international, randomized, controlled, allocation concealed, open, pragmatic, superiority clinical trial enrolling patients with ARDS. In the UK, the study received ethical approval by the London South-East Research Ethics Committee (Ref: 19/LO/1606) on 15 January 2020 with protocol version 4.0 approved on 16 June 2021. The study protocol was approved by the French Ethics Committee (Comité de Protection des Personnes Sud Mediterranee III; approved on July 30, 2020, under number2019.12.06 ter_19.11.15.76132) and Medicine Agency (Agence Nationale de Sécurité du Médicament; approved on July 3, 2020, under number 2019-A02610-57-A). The study protocol was approved by the ethics committee of Medical University of Vienna, Austria (EC No. 2056/2019) on 28 April 2020. Monitoring and oversight were provided by The Imperial Clinical Trials Unit, and independent trials steering and data monitoring and ethics committees. The study was conducted in accordance with Good Clinical Practice guidelines, local regulations, and the ethical principles described in the Declaration of Helsinki.

### Participants

The study was conducted across 3 adult intensive care units (one each in UK, France, and Austria). Adult patients were included if undergoing invasive mechanical ventilation and fulfilling ARDS criteria (as per Berlin definition: a known clinical insult with new worsening respiratory symptoms; chest radiograph with bilateral infiltrates consistent with evidence of pulmonary oedema but not fully explained by cardiac failure; Hypoxemia as defined by PaO_2_/FiO_2_ of ≤300mmHg (or ≤40kPa) (pre-ECMO PaO_2_/FiO_2_ being used should patient be placed on extracorporeal support) (14)). Exclusion criteria included age < 18 years old; the absence of an arterial catheter; decline in consent; longer than 7 days of mechanical ventilation; treatment withdrawal imminent within 24 hours; a do not attempt resuscitation order being in place, severe chronic respiratory disease requiring domiciliary ventilation and/or home oxygen therapy (except for sleep disordered breathing); requirement of veno-arterial ECMO; head trauma or other conditions with requirement of tight regulation of arterial CO_2_ level. Informed consent was obtained from the patient, personal consultee, or independent nominated professional, with retrospective consent obtained from the patient or personal consultee if possible.

#### Procedures

The Beacon Caresystem was attached to all enrolled patients, who were then randomized to having the advice of the system active (intervention) or not (control). Randomization was stratified by site, ECMO/non ECMO and COVID-19/non-COVID-19. The primary objective was to understand whether the use of the DSS changed the driving pressure applied to the patients when ventilated in a controlled-ventilation mode. Full details of study protocol have been published previously (15) and the study has been registered in clinicaltrials.gov (NCT04115709). A summary of the methods is provided here.

In the intervention arm, the DSS was attached and advice was switched on. Advice was suspended for periods in which the DSS algorithm was not able to function (i.e. ECMO, or ventilatory modes such as airway pressure release ventilation (APRV)). Advice continued until extubation, death or transfer, but was not re-initiated on re-intubation. In the control arm, the DSS was attached and advice was switched off. A detailed description for the DSS in both arms is provided in the supplemental material.

Data was collected either directly from the DSS or by input to an electronic case report form (SMART Trial, Copenhagen, Denmark).

#### Outcome measures

The primary outcome measure was average driving pressure over the period of time attached to the DSS. Secondary clinical outcome measures were. Secondary clinical outcome measures were: (1) daily average calculated delivered pressure over time, for periods of spontaneous breathing; (2) daily average calculated mechanical power over time; (3) daily average calculated oxygenation index over time; (4) daily average ventilatory ratio over time; (5) ventilator free days at 90 days; (5) time from control mode to support mode; (6) number of changes in ventilator settings per day; (7) percentage of time in control mode ventilation; (8) percentage of time in support mode ventilation; (9) total duration of mechanical ventilation; (10) tidal volume over time; (11) PEEP setting over time; (12) ventilation related complications e.g. pneumothorax and/or pneumomediastinum; (13) device malfunction event rate; (14) device related adverse event rate; and (15) number of times the advice from the Beacon system is followed through the duration of the study. All outcomes were reported from the time of randomization. The outcomes not reported in this article will be reported separately.

#### Statistical analysis

Assuming a standard deviation of 2.5 cmH2O and including a 40% dropout, 110 patients would give 90% power to detect a 2 cmH2O reduction in driving pressure. Following the onset of COVID-19, study power was re-estimated taking into consideration repeated measurements and estimating the intraclass correlation co-efficient (ICC) of driving pressure values and the coefficient of variation (CV) for days data collected (per patient) from available data. From available data, using an ICC of 0.3 and CV of 0.8, 23 patients per arm allowed for a powered analysis. Driving pressure and other repeated measurements were analyzed using a mixed model approach, including a random clustering effect per patient and fixed-effect covariates for site, ECMO and COVID status, duration of ventilation prior to DSS connection, duration of hospital admission prior to intubation, and number of days in non-ECMO and non-support-mode. Continuous variables were presented as mean (SD) or median (IQR) if non-normally distributed and appropriate log-transformations were considered where analysis residuals were non-normal. Differences between treatment groups are presented with 95% confidence interval. Categorical data is presented as number and percentage and any comparisons between the two groups were performed using the Chi-squared or Fishers exact test. Intention-to-treat basis was used for the primary analysis including all patients randomized into the study, with per protocol analysis used as follow-up. All statistical tests were 2-sided, and significance set at p < 0.05. No imputation was carried out for missing data outside of that undertaken within the mixed-effects model. Likewise, no adjustments have been made for multiple testing. Analyses were performed using SAS v9.4 software.

Per-protocol sub-group analysis was performed based on ECMO/non-ECMO status at randomization. An additional post-hoc analysis was added to consider the absence of ECMO treatment during the whole period of DSS application. This was decided to account for the extended ECMO and ventilator durations in COVID-19 patients supported on ECMO. As the study was performed during the first COVID surge, COVID-19 ECMO durations were uncertain at the time of study design.

## Results

Between 19^th^ March 2020 and 4^th^ May 2021, ninety-five patients fulfilled inclusion criteria and underwent randomization with 47 patients allocated to control and 48 patients to the intervention arm (figure 1).

**Figure 1.**
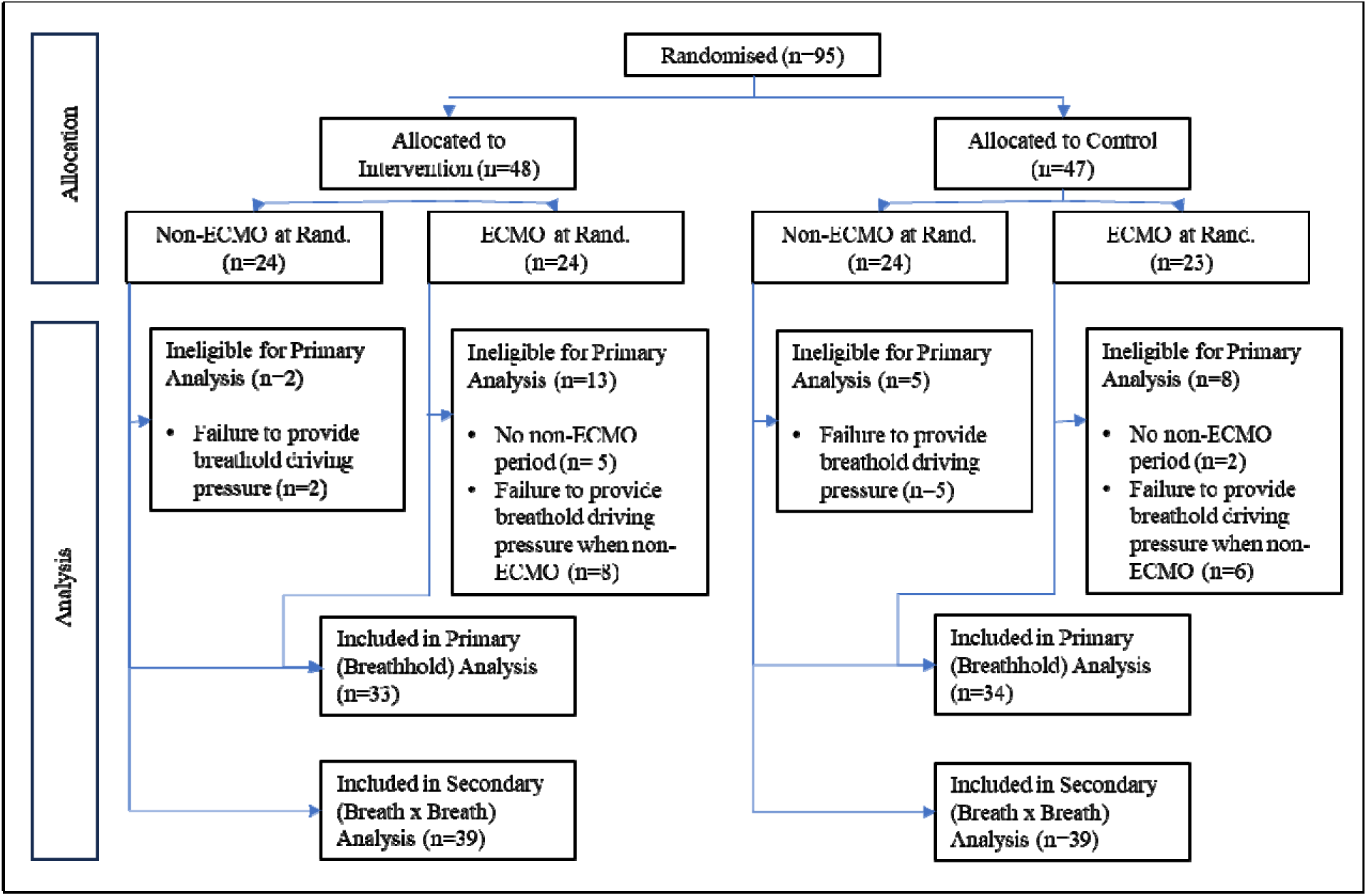
CONSORT / Flow of Patient Data

Seven patients (2 control, 5 intervention) had no periods off ECMO during their respective follow-up periods with an additional 21 patients (11 control, 10 intervention) unable to provide a breath-hold driving pressure reading due to either a) being off ECMO for a minute period (often prior to death/extubation) and/or b) being exclusively under pressure support during non-ECMO periods. As a result primary analysis was possible in 67 patients (33 intervention, 34 control). Secondary outcomes based upon continuous breath-by-breath measurements could be calculated in 78 patients.

### Patient Baseline Demographics

Demographic data by study arm are illustrated in table 1 and treatment groups were well matched for clinical parameters at baseline. Each arm had almost identical distribution of ECMO and non-ECMO patients.

**Table 1:** Patient Demographics at Randomisation.

| Variable |  | Control (N=47) | intervention (N=48) | All (N=95) |
| --- | --- | --- | --- | --- |
| Age at Consent (y) |  | 52.4 (13.58) | 53.6 (15.77) | 53.0 (14.66) |
| Ethnicity (%) | Asian | 5 (10.6) | 6 (12.5) | 11 (11.6) |
|  | Black | 7 (14.9) | 1 (2.1) | 8 (8.4) |
|  | Hispanic | 3 (6.4) | 4 (8.3) | 7 (7.4) |
|  | Indian Subcontinent | 1 (2.1) | 3 (6.3) | 4 (4.2) |
|  | Middle Eastern | 1 (2.1) | 0 | 1 (1.1) |
|  | Mixed Race | 2 (4.3) | 2 (4.2) | 4 (4.2) |
|  | White | 28 (59.6) | 32 (66.7) | 60 (63.2) |
| Gender – Male (%) | n (%) | 30 (63.8) | 32 (66.7) | 62 (65.3) |
| Height (cm) | Mean (SD) | 172.1 (9.10) | 171.0 (8.84) | 171.6 (8.94) |
| Weight (kg) | Mean (SD) | 96.4 (30.43) | 98.3 (32.83) | 97.3 (31.51) |
| BMI (kg/m<sub>2</sub>) | Mean (SD) | 32.52 (9.990) | 33.49 (10.770) | 33.01 (10.348) |
| Pred. Weight (kg) | Mean (SD) | 66.2 (10.11) | 65.3 (9.70) | 65.7 (9.86) |
| Smoking History (%) | Current smoker | 4 (8.5) | 0 | 4 (4.2) |
|  | Current vaper | 0 | 1 (2.1) | 1 (1.1) |
|  | Ex-Smoker | 7 (14.9) | 8 (16.7) | 15 (15.8) |
|  | Never Smoker | 14 (29.8) | 20 (41.7) | 34 (35.8) |
|  | Unknown | 22 (46.8) | 19 (39.6) | 41 (43.2) |
| Comorbidities – CPD (%) | n (%) | 13 (27.7) | 13 (27.1) | 26 (27.4) |
|  | Unknown | 1 (2.1) | 0 | 1 (1.1) |
| Comorbidities – CVD (%) | n (%) | 23 (48.9) | 18 (37.5) | 41 (43.2) |
|  | Unknown | 1 (2.1) | 0 | 1 (1.1) |
| Comorbidities – Metabolic & Endocrine | n (%) | 26 (55.3) | 20 (41.7) | 46 (48.4) |
|  | Unknown | 2 (4.3) | 0 | 2 (2.1) |
| Comorbidities – Chronic Immuno-suppression | n (%) | 3 (6.4) | 3 (6.3) | 6 (6.3) |
|  | Unknown | 0 | 1 (2.1) | 1 (1.1) |
| Comorbidities – Chronic neurological disease | n (%) | 2 (4.3) | 1 (2.1) | 3 (3.2) |
|  | Unknown | 7 (14.9) | 2 (4.2) | 9 (9.5) |
| Days since Ards Onset | Median [IQR] | 2 [1 – 5] | 1 [1 – 3] | 2 [1 – 4] |

### Outcomes

There was no statistically significant difference in the primary outcome variable, with values of driving pressure (table 2 and figure 2) measured from either breath hold (−0.34 cmH_2_O with intervention, 95% CI: −2.22, 1.54 cmH_2_O; p=0.72) or continuous measures (−0.23 cmH_2_O, 95% CI: −2.1, 1.67 cmH_2_O; p=0.81) being not statistically different between groups. However, there appeared to be some tendency toward tighter inter-quartile ranges in the intervention arm (figure 2).

**Figure 2.**
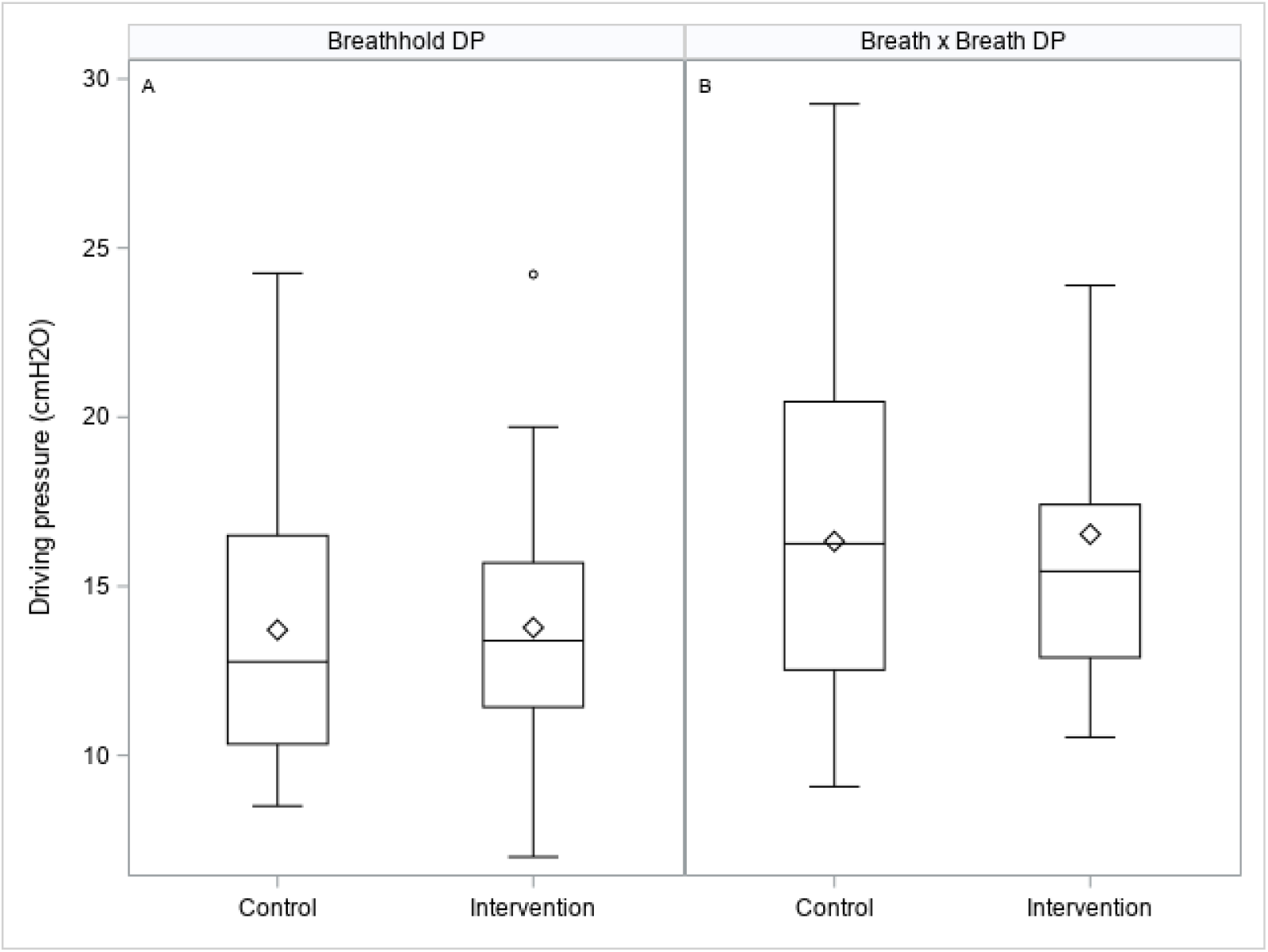
Box-plots illustrating driving pressures on control and intervention arm measured from breath holds (A) or breath by breath (B) as calculated in the ESM.

**Table 2:** Primary and secondary outcomes, all patients.

|  | Control |  | Intervention |  | Effect [95% CI] | p |
| --- | --- | --- | --- | --- | --- | --- |
|  | n | mean (SD) | n | mean (SD) |  |  |
| **Primary Outcome Measure** |  |  |  |  |  |  |
| B.Hold Driving Pressure (cmH<sub>2</sub>O) | 34 | 13.71 (4.20) | 33 | 13.78 (3.62) | -0.34 [-2.22, 1.54] | 0.72 |
| **Secondary Outcome Measures** |  |  |  |  |  |  |
| BxB Driving Pressure (cmH<sub>2</sub>O) | 39 | 16.33 (4.66) | 39 | 16.54 (4.89) | -0.23 [-2.13, 1.67] | 0.81 |
| BxB Mechanical Power (J/min) | 41 | 24.64 (8.45) | 39 | 25.80 (7.54) | -2.23 [-3.06, 2.60] | 0.87 |
| Oxygenation Index – mandatory modes | 26 | 11.10 (5.70) | 30 | 11.58 (5.61) | 0.33 [-2.34, 3.00] | 0.81 |
| Oxygenation Index – support modes | 24 | 7.69 (3.49) | 25 | 6.60 (2.39) | -1.41 [-2.74, -0.08] | 0.0370 |
| Ventilatory Ratio – mandatory modes | 26 | 2.66 (0.82) | 31 | 2.42 (0.59) | -0.31 [-0.67, 0.05] | 0.09 |
| Ventilatory Ratio – support modes | 25 | 2.55 (0.65) | 25 | 2.56 (0.72) | -0.13 [-0.47, 0.22] | 0.48 |
| Ave. Ventilator Free Days at 90 Days<sup>§</sup> | 47 | 49 [0 - 73] | 48 | 37.5 [0 - 69] | 0.0 [-20.0, 0.0] | 0.27 |
| Time from Ctrl to Support ~ | 34 | 2.35 (3.13) | 32 | 2.42 (2.98) | -0.17 [-1.58, 1.25] | 0.81 |
| Ave Daily Sett Change (PEEP)<sup>§</sup> | 44 | 1.11 [0.54-1.84] | 42 | 1.27 [0.47-2.20] | 0.14 [-0.29, 0.58] | 0.52 |
| Ave Daily Sett Change (FiO<sub>2</sub>)<sup>§</sup> | 44 | 7.50 [4.42-10.62] | 42 | 8.27 [4.32-11.7] | 1.01 [-1.07, 3.32] | 0.29 |
| Ave Daily Settings Change (PS)<sup>§</sup> | 43 | 2.65 [0.44-5.66] | 41 | 1.23 [0.24-2.70] | -0.84 [-2.10, 0.00] | 0.06 |
| Ave Daily Settings Change (PC)<sup>§</sup> | 44 | 0.66 [0 - 2.36] | 42 | 2.25 [0 - 4.86] | 0.96 [0.00, 2.15] | 0.0246 |
| Ave Daily Settings Change (RF)<sup>§</sup> | 44 | 0.94 [0 - 2.02] | 42 | 2.31 [0.95-4.96] | 1.30 [0.61, 2.30] | 0.0004 |
| % in Control Mode~ | 44 | 55.9 (36.4) | 42 | 50.3 (32.9) | -6.27 [-20.9, 8.33] | 0.40 |
| % in Support Mode~ | 44 | 31.3 (32.0) | 42 | 42.1 (31.1) | 11.6 [-1.81, 25.0] | 0.09 |
| Duration of Mech. Vent.~ | 44 | 10.52 (9.18) | 42 | 13.80 (13.40) | 3.04 [-1.35, 7.43] | 0.18 |
| Tidal Volume measured Control | 28 | 531 (113) | 33 | 520 (132) | -10.6 [-74.4, 53.1] | 0.74 |
| Tidal Volume measured Support | 27 | 644 (142) | 29 | 648 (189) | -42.82 [-125.0, 39.4] | 0.31 |
| PEEP Setting | 37 | 9.85 (2.38) | 37 | 9.88 (2.30) | 0.37 [-0.68, 1.42] | 0.49 |
| Subjects with Device related AE's<sup>§</sup> | 47 | 0 (0%) | 48 | 1 (2%) |  | 1.00 |
| Device related adverse event rate (per day)<sup>§</sup> | 47 | 0 [0 - 0] | 48 | 0 [0 - 0] | 0.0 [0.0, 0.0] | 0.32 |
| Subjects with Re-intubation events<sup>§</sup> | 47 | 2 (2%) | 48 | 2 (2%) |  | 1.00 |
| %age time disconnected prior to reintubation<sup>§</sup> | 47 | 0 [0 - 0] | 48 | 0 [0 - 0] | 0.0 [0.0, 0.0] | 0.98 |

Table showing mean (SD) or median [IQR] of Primary & Secondary outcome variables by treatment group alongside the treatment effect derived from the mixed-effects model. Model incorporates fixed-effects for COVID & Site and also adjusted for continuous variables 1) Duration of Ventilation prior to connection, 2) duration of hospitalisation prior to intubation and 3) number of days non-ECMO & non-Support and has a random effect accounting for clustering per subject.
~ No clustering effect included for these variables as data provided as an overall average per subject. \*P < 0.05, \*\*P < 0.01, \*\*\*P < 0.001
§ Median [Q1-Q3] presented. Effect represents difference in Medians is via Hodges-Lehmann estimate and distribution-free confidence interval. P-value from Wilcoxon Test
§ Frequencies presented as n (%). P-values to assess differences in event frequencies derived from Fisher's exact test.

Patients in the intervention arm had statistically improved oxygenation index when in support mode ventilation (−1.41, 95% CI: −2.76, −0.08; p=0.0370). For the subsequent post-hoc sub-group analysis of non-ECMO patients (table 3), oxygenation index was improved in the intervention arm for patients in support mode (−2.60, 95% CI:-4.13, −1.08; p=0.0010), with controlled mandatory ventilation showing a numerical improvement although did not reach statistical significance (−2.66, 95% CI −5.38, 0.06; p = 0.06). Ventilatory ratio was also significantly improved in the intervention arm for non-ECMO patients in control mode ventilation (−0.63, 95% CI: −1.08, −0.17, p= 0.0068) although this effect was reduced when extended to the full study population (−0.31, 95% CI −0.67, 0.05; p= 0.09). There is a tendency for patients in the intervention arm to spend a greater proportion of ventilator time in pressure support mode in comparison to mandatory modes (11.6%, 95% CI: −1.8, 25.0, p=0.09).

**Table 3:**
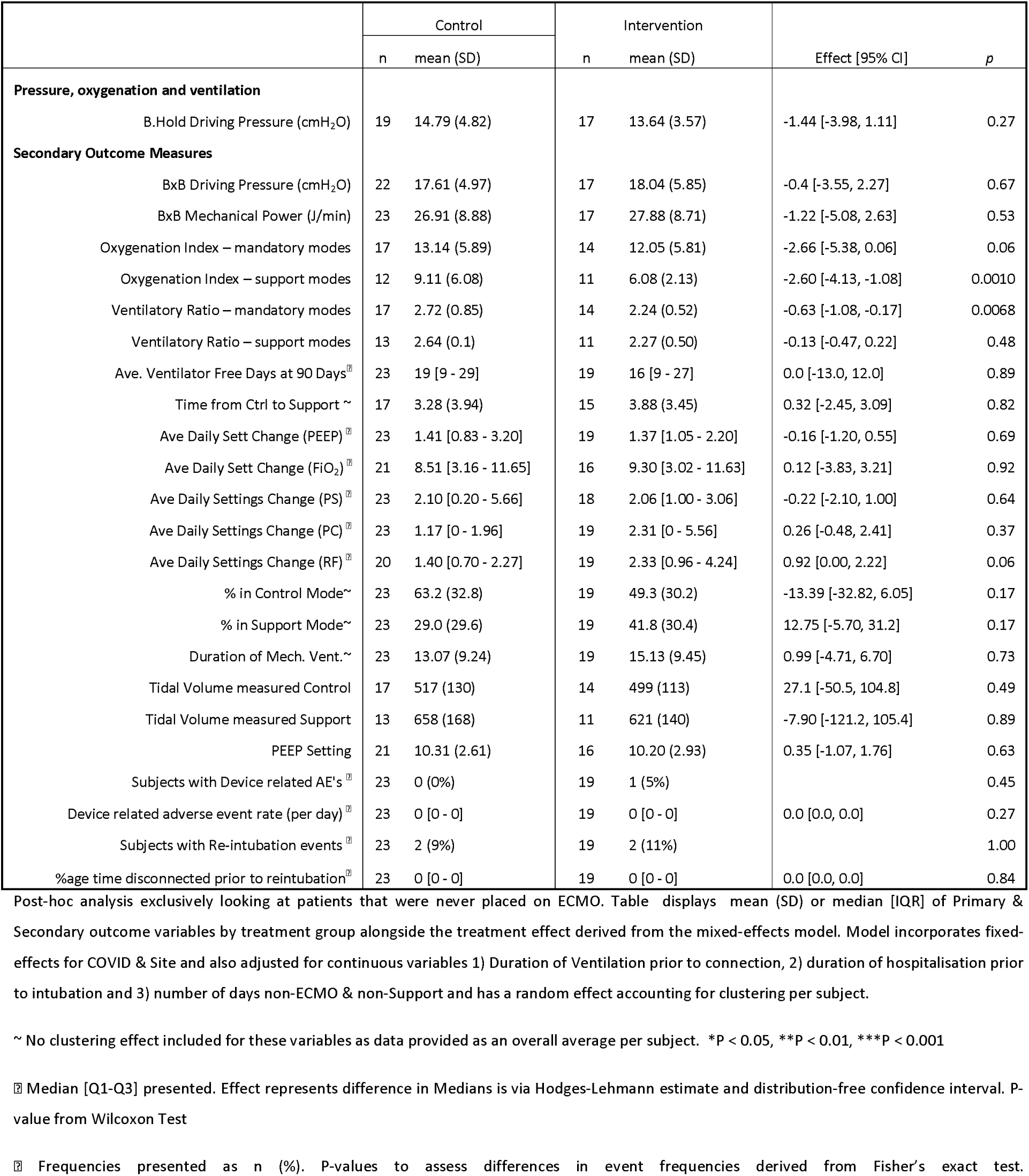
Primary and secondary outcomes, non-ECMO patients only.

Safety data is presented in Table 4 and Figure 3. No significant differences were seen in median time-to-death (control vs intervention: 19(15–59) vs 19.5(10–36)days; p=0.64). Under the full trial population there was a numerical, but not statistically significant, increase in mortality in the intervention arm (38% vs 26%) which was not present in non-ECMO patients (37% vs 39%). There were no differences between ventilation related complications, device malfunction rates or device related adverse events. No significant difference was seen in the time spent or incidence of hypoxaemia or hypercapnia in patients ventilated on control mode between groups. In support mode, in terms of per-patient percentage of their respective DSS application, significantly less time was spent at very low respiratory rates (< 12 breaths per minute) in the intervention arm (difference in medians −0.2%, 95% CI −1.6, 0.0; p= 0.0167), and at high CO_2_ levels (>7 kPa), although this was not statistically significant (difference in medians −0.0%, 95% CI −1.1, 0.0; p = 0.06), figure 3, table 4.

**Figure 3.**
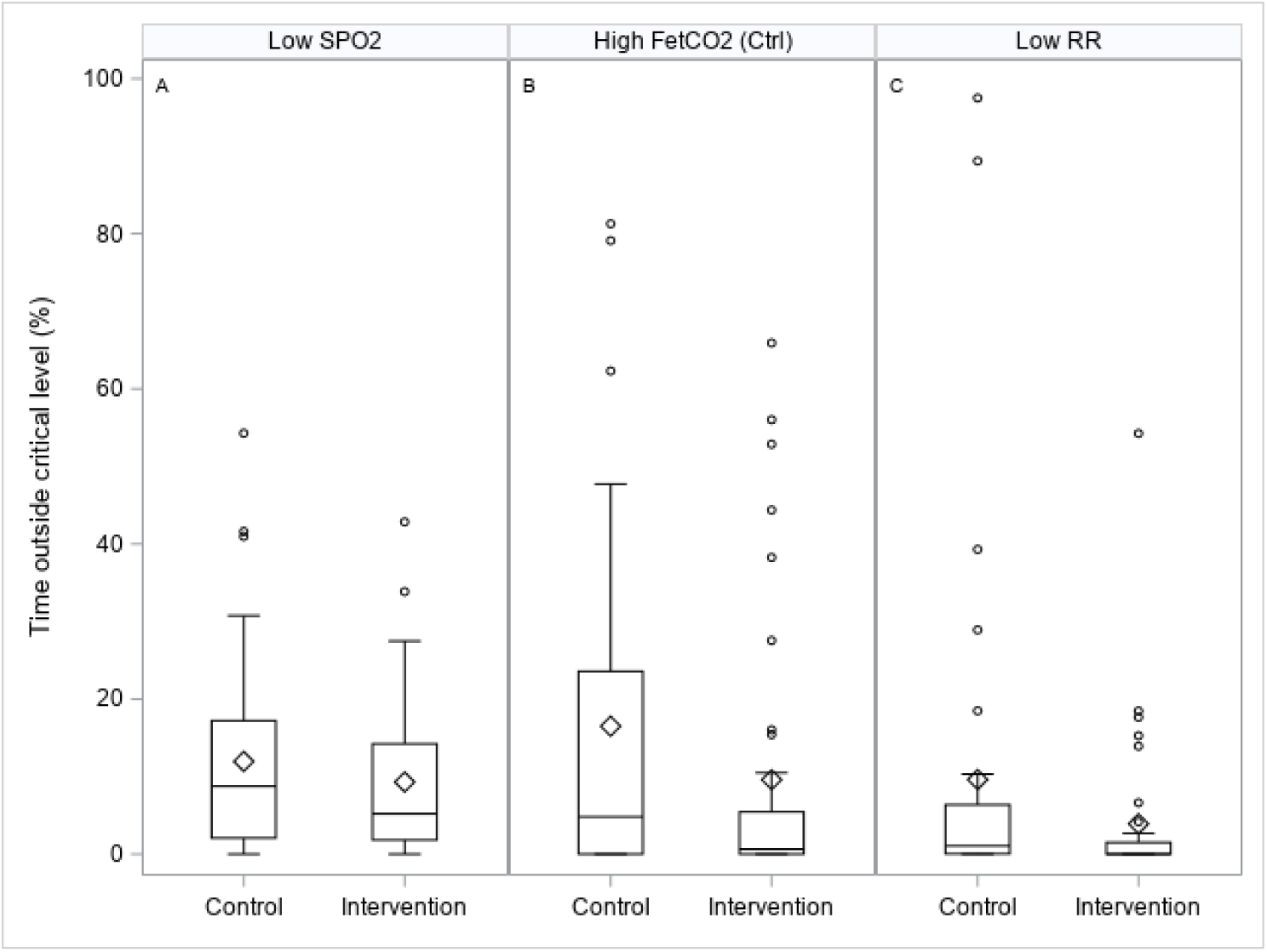
Box-plots illustrating safety measures for the percentage time spent: A - below SpO_2_ values of 88%; B - above end tidal CO_2_ values of 7 kPa in control ventilation; and C - below respiratory rate of 12 breaths per minute in pressure support ventilation.

**Table 4.** Safety analysis showing 90-day mortialiy and key ventilation parameters.

|  | Control |  |  | Intervention |  |  | Comparison |  |
| --- | --- | --- | --- | --- | --- | --- | --- | --- |
|  |  | Deaths | Days to event |  | Deaths | Days to event |  |  |
|  | n | n(%) | Med. [Q1-Q3] | n | n(%) | Med. [Q1-Q3] | Δ [95% CI] | p-value |
| Death – all patients | 47 | 12 (26) | 19.0 [15-59] | 48 | 18 (38) | 19.5 [10-36] | -1.5 [-18.0, 14.0] | 0.64 |
| Death – non-ECMO patients | 23 | 9 (39) | 19.0 [18-22] | 19 | 7 (37) | 16.0 [6-21] | -6.0 [-44.0, 5.0] | 0.20 |
|  | n | Mean | Median | n | Mean | Median | Δ [95% CI] | p-value |
| % time below SpO<sub>2</sub> 88% | 44 | 12.0 | 8.7 | 42 | 9.3 | 5.2 | -0.98 [-5.07, 1.51] | 0.44 |
| % time FetCO<sub>2</sub> > 7 kPa, mandatory | 32 | 16.5 | 4.8 | 38 | 9.6 | 0.6 | -0.05 [-7.96, 0.00] | 0.19 |
| % time FetCO<sub>2</sub> > 7 kPa, support | 32 | 16.7 | 0.1 | 34 | 7.4 | 0.0 | 0.00 [-1.06, 0.00] | 0.06 |
| % time RR < 12, support | 34 | 9.6 | 1.1 | 35 | 3.9 | 0.0 | -0.20 [-1.58, 0.00] * | 0.0167 |
| % time RSBi >100, support | 32 | 1.0 | 0.0 | 34 | 0.8 | 0.0 | 0.00 [0.00, 0.00] | 0.78 |

Δ [95% CI] represents difference in medians and a distribution-free 95% CI via Hodges-Lehmann estimate and distribution-free confidence interval.
P-value from Wilcoxon Test. \*P < 0.05, \*\*P < 0.01, \*\*\*P < 0.001

The application of the DSS resulted in a significantly increased number of ventilator changes for pressure settings during control mode ventilation (control vs intervention: 0.65 vs 2.25 changes per day; P<0.05) and respiratory frequency (control vs intervention: 0.94 vs 2.31 changes per day; P<0.001). Table 5 illustrates the uptake of advice over a two-hour window following advice presentation, where advice was to change ventilator settings to those different from current values. Indeed, only 44% of advice given to change pressure support and 66% for FIO_2_ was actioned. Advice was often not followed, meaning that no ventilator changes were made during the two-hour window following advice presentation. Of note, changes in ventilator settings in the opposite direction to advice presented within the two-hour window were minimal.

**Table 5.** application of system’s advice - intervention arm only.

|  |  | Advice Followed (%) | Advice Ignored (%) | Changes made which were opposite to the advice suggested (%) |
| --- | --- | --- | --- | --- |
|  | n | Median [Q1-Q3] | Median [Q1-Q3] | Median [Q1-Q3 ] |
| FiO<sub>2</sub> , PEEP | 34 | 65.7 [40.0-72.7] | 25.0 [12.5-40.0] | 12.1 [0.0 -0.0] |
| PC, RR (mandatory) | 19 | 60.0 [0.0-71.4] | 33.3 [13.3-83.3] | 0.0 [0.0-14.3] |
| VT, RR (mandatory) | 7 | 61.5 [14.3-83.3] | 50.0 [9.5-66.5] | 16.7 [4.8-50.0] |
| Pressure support | 21 | 43.8 [0-60.0] | 39.6 [0.0-54.5] | 0.0 [0.0-10.0] |

Table presenting medians and quartiles of how advice was actioned following Beacon recommendations. Advice recommended by Beacon was either followed, ignored (with no change made to ventilator settings) or different to what the clinician actioned.

No other significant differences were seen in secondary endpoints assessing physiological measurements or ventilation duration.

## Discussion

This study was designed to evaluate this decision support system in patients with ARDS receiving mechanical ventilation prior to COVID onset and repurposed for implementation during the pandemic under extremely challenging circumstances, with inclusion of patients with ARDS from both COVID and non-COVID etiologies.

No significant differences were seen in driving pressure, the primary outcome of the study; however, patients in the intervention arm tended to have tighter regulation of driving pressure as compared to those in standard care. This may be important as it is perhaps more crucial to reduce the incidence of high levels of driving pressure than to modify the median value delivered as values below 15cmH_2_O may reflect little increased risk for the patient (16).

Application of DSS advice showed optimization of physiological state under certain conditions. Significant improvement in oxygenation index was seen without increased incidence of hypoxaemia, consistent with an appropriate use of FiO_2_. A significantly improved ventilatory ratio was seen for non-ECMO patients under controlled ventilation modes. There were a greater number of changes in pressure control and respiratory frequency in the intervention group with a reduced, yet not statistically significant, number of changes required in pressure support settings. Support mode ventilation was delivered at significantly reduced time spent with a respiratory rate less than 12 breath/min, values below which have been shown previously to be associated with ventilatory over assistance (17). Furthermore, there was a trend towards a reduction in the percentage time spent at end tidal carbon dioxide levels greater than 7kPa (p = 0.06), alongside, non-significant tendencies for pressure support reduction of about 1-2cmH_2_O. No significant detrimental effects were observed by application of the advice.

Despite the difficulties of implementing a complex, “open loop” DSS intervention during pandemic conditions, the data shows advice was applied about 60% of the time, with the exception of advice on pressure settings for which advice was applied a median of 44%. However, despite a significant proportion of advice was either ignored or not seen, optimization of gas exchange still occurred. This may reflect the system requesting small changes in ventilatory parameters which may be at odds to purely clinician made changes. This lack of adoption may be enhanced during the pandemic surges and contrasts with other studies evaluating the application of the system in short controlled periods (9). Importantly, for advice on pressure settings in either mandatory or support modes, clinicians tended not to make changes different to the advice (as represented by a median change count of zero within both mode settings). For FiO_2_, clinicians made settings in the opposite direction a median of 12% of the time. The lack of implementation of a higher percentage of advice places limitations on the interpretation of these results, illustrating that improvements may be made when the advice is used as an augmentation of current care rather than a replacement. If such a system were to be used in a closed loop context, analysis of the reasons for difference in opinions would be necessary. Nonetheless, this system also enables an understanding of the frequency of ventilator changes that are made and the implementation space for such a device.

Other decision support systems and closed-loop control systems for mechanical ventilation have been evaluated in prospective studies, but few in relation to the management of ARDS. Of note, East et. al. showed physiological efficacy through significant improvement in morbidity scores in 200 ARDS patients randomized to decision support advice based on empirical rules rather than physiological models in a non-commercial system (18). The most widely applied and evaluated routine commercial tools - SmartCare (Dräger Medical) and Intelli-Vent Adaptive Support Ventilation (INTELLiVENT-ASV) or its predecessor ASV (Hamilton Medical), apply closed-loop automation rather than open loop advice. SmartCare provides control during support mode ventilation and has been shown in some studies to significantly reduce weaning duration in patients after evaluation of weaning readiness (19, 20). ASV and INTELLiVENT-ASV control the patient through all phases of ventilation and studies have shown significant reductions in weaning time (21, 22) or total ventilator time (23, 24). However, these have been in fast track cardiac patients or patients with COPD, but excluded patients with ARDS. A single retrospective study with Intelli-Vent ASV has shown significant reduction in driving pressure in 51 COVID-ARDS patients two hours after conversion to INTELLiVENT-ASV suggesting potential for improvement in care without evidence provided by prospective evaluation (25). Hence, such novel technologies using personalized approaches may lead to improvements in weaning (26).

Titration of ventilator setting in the acute phase based upon pressure-volume curves and applying optimal PEEP and driving pressure settings are associated with better outcomes in ARDS (27, 28), however, it remains uncertain whether this benefit is due to optimized PEEP or use of low tidal volume ventilation (29). Indeed, within the current era of ARDS with lung protective ventilation strategies as a standard of care in the specialist centre involved in this study, the impact on driving pressure may not have been modulatable. Furthermore, the different pathophysiology of COVID-ARDS versus ARDS of conventional non-COVID aetiologies may have played a role in how the system could impact driving pressure versus more composite measure such as oxygenation index and ventilratory ratio (30–33).

There are several limitations of this study. The primary was the challenging clinical environment during COVID pandemic conditions, which was not ideal for evaluation and implementation of such a new device. This, we believe, explains the high percentage of advice ignored. In addition, two sites were ECMO centres with many enrolled patients requiring ECMO support and the device being unable to give advice during the ECMO period led to greater heterogeneity in the patient cohort. Evaluation of differences in driving pressure in those patients who often had a few days non-ECMO ventilation, followed by a prolonged ECMO period, and then a further period of non-ECMO ventilation, inevitably resulted in comparisons which might be biased by ECMO duration. Indeed, our sensivity analysis showed that improvements in physiological status were more pronounced in the non-ECMO group.

## Conclusions

This study is the first evaluation of a physiological model-based decision support system for advice on mechanical ventilation in patients with COVID-19 and non-COVID-19 ARDS. Application of about 60% of advice improved physiological state, but with no significant difference in driving pressure as a primary outcome measure. There was greater homogeneity in ventilatory management with the system being safe to implement in patients with ARDS and during pandemic conditions. Further clinical effectiveness studies examining the implementation of such a DSS in ARDS could assist in assessing if such interventions show any clinical significance and/or cost benefits.

## Data Availability

All data produced in the present study are available upon reasonable request to the authors

## References

1. Panwar R, Madotto F, Laffey JG, Haren FMP van. Compliance Phenotypes in Early Acute Respiratory Distress Syndrome before the COVID-19 Pandemic. Am J Respir Crit Care Med 2020;202:1244–1252.

2. Karbing DS, Panigada M, Bottino N, Spinelli E, Protti A, Rees SE, et al. Changes in shunt, ventilation/perfusion mismatch, and lung aeration with PEEP in patients with ARDS: a prospective single-arm interventional study. Crit Care 2020;24:111.

3. Gattinoni L, Coppola S, Cressoni M, Busana M, Rossi S, Chiumello D. Covid-19 Does Not Lead to a “Typical” Acute Respiratory Distress Syndrome. Am J Respir Crit Care Med 2020;0:1299–1300.

4. Scaramuzzo G, Karbing DS, Fogagnolo A, Mauri T, Spinelli E, Mari M, et al. Heterogeneity of Ventilation/Perfusion Mismatch at Different Levels of PEEP and in Mechanical Phenotypes of COVID-19 ARDS. Respir care 2022;68:188–198.

5. Pelosi P, Ball L, Barbas CSV, Bellomo R, Burns KEA, Einav S, et al. Personalized mechanical ventilation in acute respiratory distress syndrome. Crit Care 2021;25:250.

6. Patel BV, Haar S, Handslip R, Auepanwiriyakul C, Lee TM-L, Patel S, et al. Natural history, trajectory, and management of mechanically ventilated COVID-19 patients in the United Kingdom. Intens Care Med 2021;1–17.doi:10.1007/s00134-021-06389-z.

7. Constantin J-M, Jabaudon M, Lefrant J-Y, Jaber S, Quenot J-P, Langeron O, et al. Personalised mechanical ventilation tailored to lung morphology versus low positive end-expiratory pressure for patients with acute respiratory distress syndrome in France (the LIVE study): a multicentre, single-blind, randomised controlled trial. The lancet Respiratory medicine 2019;7:870–880.

8. Calfee CS, Delucchi K, Parsons PE, Thompson BT, Ware LB, Matthay MA, et al. Subphenotypes in acute respiratory distress syndrome: latent class analysis of data from two randomised controlled trials. Lancet Respir Med 2014;2:611–620.

9. Morris AH, Stagg B, Lanspa M, Orme J, Clemmer TP, Weaver LK, et al. Enabling a learning healthcare system with automated computer protocols that produce replicable and personalized clinician actions. J Am Méd Inform Assoc 2021;28:1330–1344.

10. Needham DM, Colantuoni E, Mendez-Tellez PA, Dinglas VD, Sevransky JE, Himmelfarb CRD, et al. Lung protective mechanical ventilation and two year survival in patients with acute lung injury: prospective cohort study. BMJ 2012;344:e2124.

11. Rees SE, Karbing DS. Determining the appropriate model complexity for patient-specific advice on mechanical ventilation. Biomed Eng Biomedizinische Technik 2017;62:183–198.

12. Karbing DS, Spadaro S, Dey N, Ragazzi R, Marangoni E, Corte FD, et al. An Open-Loop, Physiologic Model–Based Decision Support System Can Provide Appropriate Ventilator Settings. Crit Care Med 2018;46:e642–e648.

13. Spadaro S, Karbing DS, Corte FD, Mauri T, Moro F, Gioia A, et al. An open-loop, physiological model based decision support system can reduce pressure support while acting to preserve respiratory muscle function. J Crit Care 2018;48:407–413.

14. FORCE TADT. Acute Respiratory Distress Syndrome: The Berlin Definition. Jama 2012;307:2526–2533.

15. Patel B, Mumby S, Johnson N, Falaschetti E, Hansen J, Adcock I, et al. Decision support system to evaluate ventilation in the acute respiratory distress syndrome (DeVENT study)—trial protocol. Trials 2022;23:47.

16. Amato MBP, Meade MO, Slutsky AS, Brochard L, Costa ELV, Schoenfeld DA, et al. Driving Pressure and Survival in the Acute Respiratory Distress Syndrome. New Engl J Medicine 2015;372:747–755.

17. Pletsch-Assuncao R, Pereira MC, Ferreira JG, Cardenas LZ, Albuquerque ALP de, Carvalho CRR de, et al. Accuracy of Invasive and Noninvasive Parameters for Diagnosing Ventilatory Overassistance During Pressure Support Ventilation*. Crit Care Med 2018;46:411–417.

18. East TD, Heermann LK, Bradshaw RL, Lugo A, Sailors RM, Ershler L, et al. Efficacy of computerized decision support for mechanical ventilation: results of a prospective multi-center randomized trial. Proc AMIA Symp 1999;251–5.

19. Lellouche F, Mancebo J, Jolliet P, Roeseler J, Schortgen F, Dojat M, et al. A Multicenter Randomized Trial of Computer-driven Protocolized Weaning from Mechanical Ventilation. Am J Respir Crit care Med 2006;174:894–900.

20. Burns KEA, Meade MO, Lessard MR, Hand L, Zhou Q, Keenan SP, et al. Wean Earlier and Automatically with New Technology (the WEAN Study). A Multicenter, Pilot Randomized Controlled Trial. Am J Respir Crit Care Med 2013;187:1203–1211.

21. Sulzer CF, Chioléro R, Chassot P-G, Mueller XM, Revelly J-P. Adaptive Support Ventilation for Fast Tracheal Extubation after Cardiac Surgery. Anesthesiology 2001;95:1339–1345.

22. Gruber PC, Gomersall CD, Leung P, Joynt GM, Ng SK, Ho K, et al. Randomized Controlled Trial Comparing Adaptive-support Ventilation with Pressure-regulated Volume-controlled Ventilation with Automode in Weaning Patients after Cardiac Surgery. Anesthesiology 2008;109:81–87.

23. Kirakli C, Naz I, Ediboglu O, Tatar D, Budak A, Tellioglu E. A Randomized Controlled Trial Comparing the Ventilation Duration Between Adaptive Support Ventilation and Pressure Assist/Control Ventilation in Medical Patients in the ICU. Chest 2015;147:1503–1509.

24. Kirakli C, Ozdemir I, Ucar ZZ, Cimen P, Kepil S, Ozkan SA. Adaptive support ventilation for faster weaning in COPD: a randomised controlled trial. Eur Respir J 2011;38:774–80.

25. Buiteman-Kruizinga LA, Mkadmi HE, Neto AS, Kruizinga MD, Botta M, Schultz MJ, et al. Effect of INTELLiVENT-ASV versus Conventional Ventilation on Ventilation Intensity in Patients with COVID-19 ARDS—An Observational Study. J Clin Med 2021;10:5409.

26. Rose L, Schultz MJ, Cardwell CR, Jouvet P, McAuley DF, Blackwood B. Automated versus non-automated weaning for reducing the duration of mechanical ventilation for critically ill adults and children: a cochrane systematic review and meta-analysis. Crit Care 2015;19:48.

27. Amato MB, Barbas CS, Medeiros DM, Schettino G de P, Filho GL, Kairalla RA, et al. Beneficial effects of the “open lung approach” with low distending pressures in acute respiratory distress syndrome. A prospective randomized study on mechanical ventilation. Am J Respir Crit Care Med 1995;152:1835–1846.

28. Amato MBP, Barbas CSV, Medeiros DM, Magaldi RB, Schettino GP, Lorenzi-Filho G, et al. Effect of a Protective-Ventilation Strategy on Mortality in the Acute Respiratory Distress Syndrome. N Engl J Med 1998;338:347–354.

29. Network ARDS, Brower RG, Matthay MA, Morris A, Schoenfeld D, Thompson BT, et al. Ventilation with Lower Tidal Volumes as Compared with Traditional Tidal Volumes for Acute Lung Injury and the Acute Respiratory Distress Syndrome. N Engl J Med 2000;342:1301–1308.

30. Patel BV, Arachchillage DJ, Ridge CA, Bianchi P, Doyle JF, Garfield B, et al. Pulmonary Angiopathy in Severe COVID-19: Physiologic, Imaging and Hematologic Observations. Am J Resp Crit Care 2020;doi:10.1164/rccm.202004-1412oc.

31. Tsolaki VS, Zakynthinos GE, Mantzarlis KD, Deskata KV, Papadonta M-EE, Gerovasileiou ES, et al. Driving Pressure in COVID-19 Acute Respiratory Distress Syndrome Is Associated with Respiratory Distress Duration before Intubation. Am J Respir Crit Care Med 2021;204:478–481.

32. Pozzi T, Collino F, Brusatori S, Romitti F, Busana M, Moerer O, et al. Specific Respiratory System Compliance in COVID-19 and Non–COVID-19 Acute Respiratory Distress Syndrome. Am J Respir Crit Care Med 2023;208:328–330.

33. Bos LDJ, Sjoding M, Sinha P, Bhavani SV, Lyons PG, Bewley AF, et al. Longitudinal respiratory subphenotypes in patients with COVID-19-related acute respiratory distress syndrome: results from three observational cohorts. Lancet Respir Med 2021;9:1377–1386.

