## Supplementary Materials for "Decision support system to evaluate VENTilation in the Acute Respiratory Distress Syndrome"

**Title: De**cision support system to evaluate **VENT**ilation in the Acute Respiratory Distress Syndrome

**see acknowledgements

Index of supplementary material

S.1) Structure and Function of Beacon Caresystem

S.2) Study outcome calculations.

S.3) Data collection from the Beacon Caresystem

S.4) Evaluation of advice use

S.5) References

S.6) Acknowledgements

**S.1) Structure and Function of Beacon Caresystem**

This section describes the structure of the Beacon Caresystem. Included is a brief description of mathematical models and the function of the system in providing advice on ventilator settings. Much of this supplementary material has been provided previously by the some of the authors of this work [1,2,3].

Mathematical models included in the CDSS

Figure S.1.1 illustrates the structure of the mathematical models, a full description having been published previously [4]. The system includes models of pulmonary gas exchange; respiratory mechanics; acid-base chemistry of blood, interstitial fluid, tissues and cerebral spinal fluid; respiratory drive and ventilation. In addition, the models include the effects of PEEP on gas exchange, pulmonary mechanics, and ventilation. Models are tuned to the individual patient's data through measurement of respiratory gas flows and pressures; calorimetry and capnography measurements of respiratory gas fractions of $O_2$ and $CO_2$, and subsequent calculation of oxygen utilisation ($VO_2$) and carbon dioxide production ($VCO_2$); pulse oximetry measurement of arterial oxygen saturation; and arterial blood measurements of acid-base, oxygenation and haemoglobin fractions. The model of pulmonary gas exchange is tuned to the appropriate matching of ventilation and perfusion to account for $O_2$ and $CO_2$ differences between arterial and end tidal gas values. To do so an arterial blood gas (ABG) is required on system start up. In some patients, the system also requires modification of $FIO_2$ to two levels for 2-5 minutes at each level to tune the pulmonary gas exchange model to the patient [5,6,7]. The respiratory mechanics model is tuned to dynamic compliance. The model of acid-base chemistry of the blood is tuned to measured values of arterial pH, $PCO_2$, $PO_2$, and $SO_2$, and haemoglobin concentration, with the acid-base chemistry of the cerebrospinal fluid (CSF) regulated to arterial bicarbonate values to account for chemical changes in respiratory drive. The respiratory drive model is tuned to describe the relationship between alveolar ventilation (VA) and arterial acid-base and oxygenation status. The ventilation model is tuned to the measured serial dead space (Vd) to allow calculation of alveolar ventilation. A series of algorithms are present to re-tune the models as new measurements present, or if the patient condition changes. These models are used to simulate the effect of changes in ventilator settings, with the system generating advice based upon simulated values. The mathematical models describing the response to changes in PEEP on pulmonary gas exchange and mechanics has been described in previous supplementary material [2].

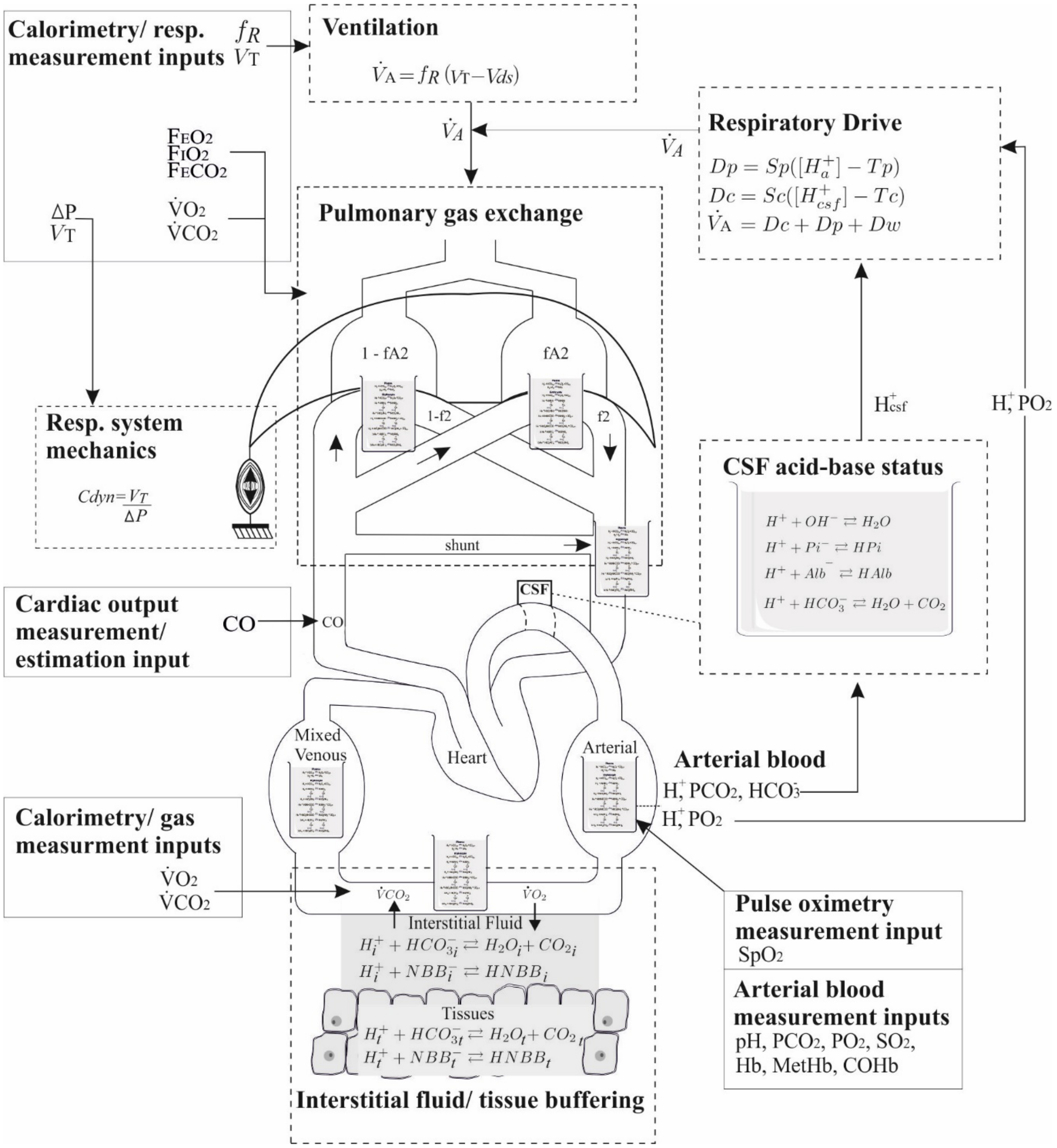


Figure S.1.1: Mathematical models included in the system, modified from [4]. Mathematical models are indicated by dashed boxes with the exception of the acid-base model of blood, which is illustrated as a beaker of blood in each blood compartment where it is used. Measured input values are indicated by solid boxes.

*Ventilator advice*

Advice is calculated by the system using the mathematical models tuned to the specific patient measurements, along with a set of clinical preference functions. These quantify and balance the negative outcomes of mechanical ventilation in a decision theoretic approach [8]. Model tuning, the clinical balances and the resulting advice screen are illustrated in figure S.1.2 as published previously [9]. The balances are between: over and under oxygenation; the risk of ventilator induced lung injury due to high pressures, respiratory rate or tidal volumes against the risk of acidosis with insufficient $CO_2$ expiration; and for pressure support, the risk of respiratory muscle atrophy with over support against a stressful situation for the patient with excessive work of breathing for under support. Advice is provided in steps moving the patient toward ventilator settings which the system calculates as providing the minimum risk for the patient at that point in time. Following change of ventilator settings by the clinician, a period of 5–20 min is waited by the system to ensure the full effects of the ventilator changes are complete, and new advice is generated. If system calculations show no further changes can improve patient state then no further advice is given until patient state changes. In this way, the frequency of advice depends upon the stability of the patient and the difference between current and 'optimal' ventilator settings. Physiological models are re-tuned as the patient state changes and on measured response to changes in ventilator settings.

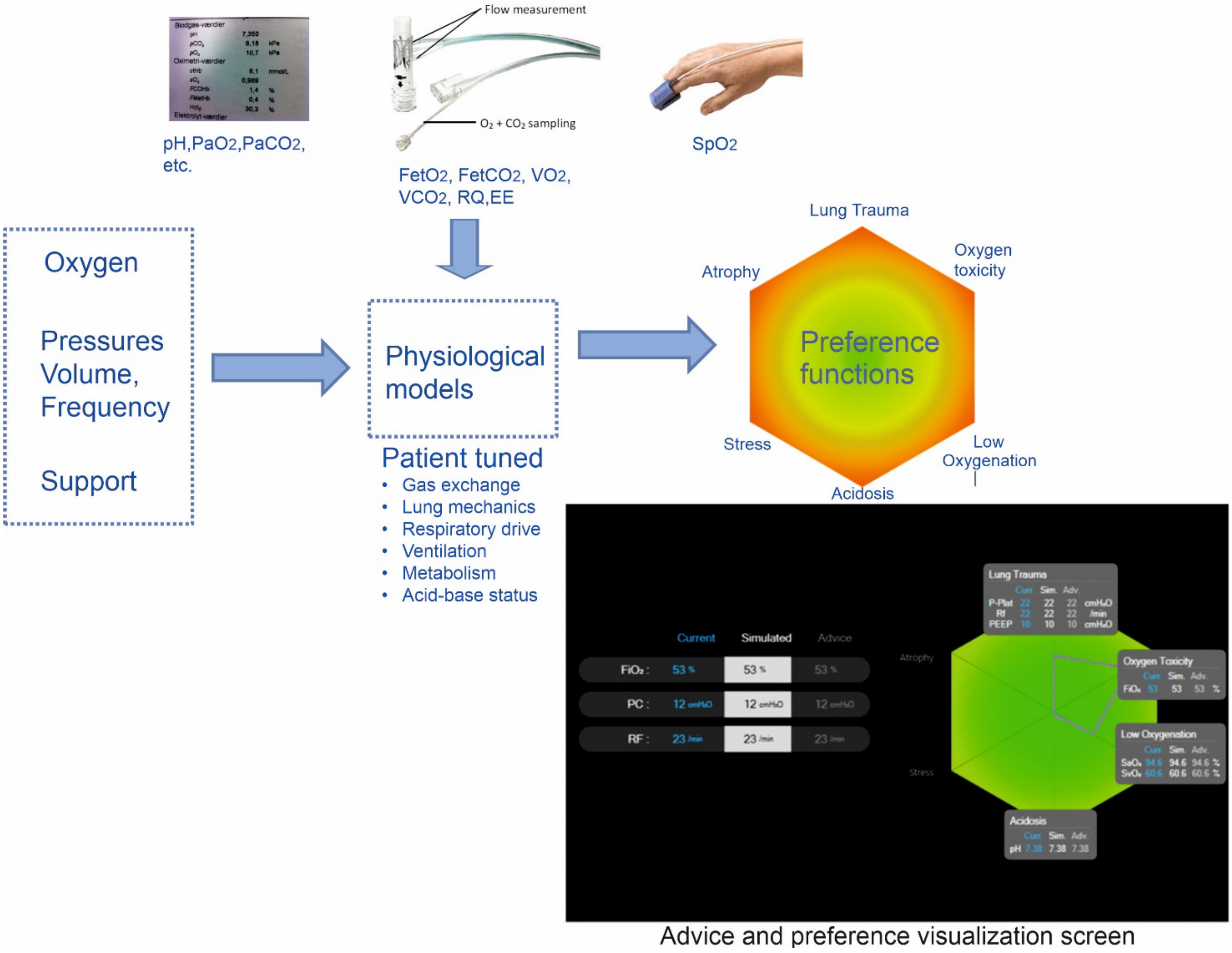


Figure S.1.2 – Beacon Caresystem structure and advice screen, see [9] for details. Modified from [9].

**S.2) Study outcome calculations**

The following text described the calculation of study outcomes where not directly measured.

Primary outcome

Driving pressure. For all patients, continuous measurements of pressure were measured at the t-tube by the Beacon Caresystem. Driving pressure was calculated as illustrated in figure S.2.1. For breaths including an inspiratory pause, plateau pressure was calculated from the last second of the pause period, and driving pressure calculated as the difference between this and PEEP identified from the eventual expiratory plateau.

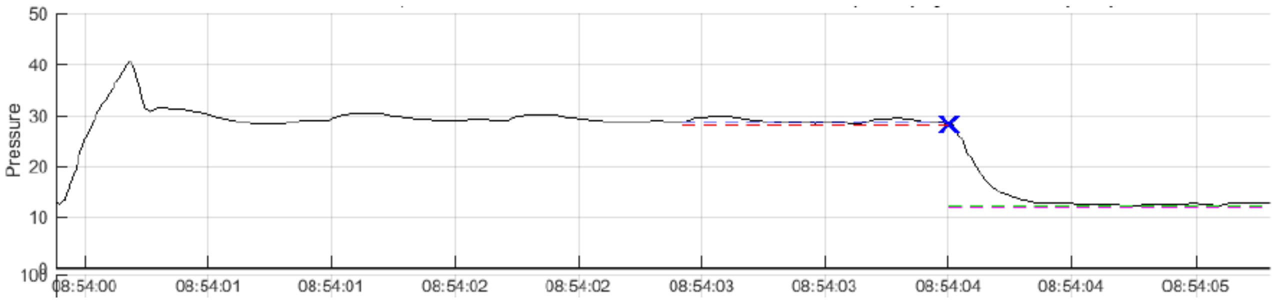


Figure S.2.1 – an example of calculation of driving pressure.

Secondary outcomes

Breath by Breath Driving pressure. A value of driving pressure was also calculated for every breath in mandatory mode in the same way as illustrated in figure S.2.1. This was performed to allow comparison over al breaths between study groups, with driving pressure values typically higher than those from inspiratory pauses due to incomplete equilibrium amongst lung compartments with different time constants.

Breath by Breath mechanical power: For each value of driving pressure values were calculated for mechanical power (MP) using the simplified form of Gattinoni et al (10) , i.e.

$$MP = 0.098 * RR * Vt * (Peak - (0.5*(Pplat-PEEP)))$$

To do so the corresponding Peak pressure was identified from the pressure waveform. In addition, flow waveforms were integrated to obtain tidal volume (Vt).

Oxygenation index: A value of the oxygenation index was calculated according to the following formula

$$\text{Oxygenation index} = \frac{F_IO_2 \text{ x } P_{AW}}{P_aO_2}$$

Where mean airway pressure (PAW) was calculated for volume and pressure modes according to Hess (11) as

PAW = 0.5*(PIP - PEEP)*(TI/Ttot) + PEEP : For volume modes, and

PAW = (PIP - PEEP)*(TI/Ttot) + PEEP : For pressure modes

Values were calculated at each morning on measurement of $PaO_2$ from the first morning blood sample, with all other data recorded from the Beacon Caresystem at that time. Average values were calculated per patient for both mandatory and support mode ventilation.

Ventilatory ratio:

Ventilatory ratio was calculated using the following formula

$$\text{Ventilatory Ratio} = \frac{minute\ ventilation\ \left(\frac{ml}{min}\right) * \mathrm{P_aCO_2}\ (\mathrm{mmHg})}{\text{predicted body weight(kg)}*100*37.5}$$

Values were calculated at each morning on measurement of $PaCO_2$ from the first morning blood sample, with all other data recorded from the Beacon Caresystem at that time, apart from predicted body weight which was recorded for each patient at the start of the protocol. Average values were calculated per patient for both mandatory and support mode ventilation.

Average ventilator free days and 90 Days: (I don't have this in my notes

Time from control mode to support mode: All changes in mode were recorded automatically by the Beacon Caresystem. A change from control to support mode was defined as a mode change to support mode which was maintained for a period of more than 30 minutes.

Number of changes in ventilator settings per day: All changes in ventilator settings were recorded automatically by the Beacon Caresystem, and cumulative counts obtained per day pr setting.

% of time in control mode ventilation: Mode was automatically collected per minute for the complete duration by the Beacon Caresystem, allowing calculation of the total and percentage time spent in control mode.

% of time in support mode ventilation: Mode was automatically collected per minute for the complete duration by the Beacon Caresystem, allowing calculation of the total and percentage time spent in support mode.

Duration of mechanical ventilation: Values of intubation and extubation times were recorded in the electronic case report form.

Tidal volume measured, Control: Average minute by minute values of tidal volume were recorded automatically by the Beacon Caresystem and used to calculate the average tidal volume used per patient when in mandatory modes.

Tidal volume measured, Support: Average minute by minute values of tidal volume were recorded automatically by the Beacon Caresystem and used to calculate the average tidal volume used per patient when in spontaneous modes.

PEEP setting: Average values of PEEP set on the ventilator were recorded automatically by the Beacon Caresystem.

Device related adverse event rate (per day): Adverse events were recorded in the electronic case report form as specified in the trial protocol (1).

Subjects with re-intubation events: Re-intubation was recorded in the electronic case report form.

In addition to the secondary outcomes described above, patient mortality was recoded in the electronic case report form, and an analysis of safety performed from continuous measurements recorded by the Beacon Caresystem. This included:

Oxygenation safety: An analysis of the duration of time spent with an SpO2 < 88% was recorded per patient.

Hypercapnia safety: An analysis of the duration of time spent with an end tidal $CO_2$ level > 7 kPa was recorded per patient.

Ventilation over-support: An analysis was performed of the time spent with a respiratory frequency lower than 12 breaths per minute in support mode ventilation, where 12 breath per minute has previously been indicated as a threshold for over support (12).

Ventilation under-support: An analysis was performed of the time spent with a rapid shallow breathing index > 100 breaths/min L, indicative of under-support.

**S.3) Data collection from the Beacon Caresystem.**

The system includes a tablet computer, plus a gas module including volumetric capnography and indirect calorimetry, with both mounted on the ventilator or a separate trolley. The patient is connected to the system via a respiratory tube measuring flow and with side stream sampling for measurement of $O_2$ and $CO_2$. The system is also connected to the ventilator data output to allow for recoding of ventilation settings and changes and ventilator mode.

The following physiologic data were collected from the measurements taken by the BC, and saved as averages over a 1 minute duration for the full duration. Data was saved as spreadsheets including 1440 rows, one per minute, for each day of ventilation.

$SpO_2$ – Pulse oximetry oxygen saturation (%).
$FetO_2$ – End tidal oxygen fraction.
$FetCO_2$ – End tidal CO2 fration.
$VO_2$ - Oxygen consumption (l/min)
$VCO_2$ – Carbon dioxide production (l/min)
Rf – measured respiratory frequency.
Vt – measured tidal volume (l)
Vd – Anatomical dead space estimated from volumetric capnography (l).
Pplat – Measured plateau pressure ($cmH_2O$).
Comp – Respiratory system compliance ($l/cmH_2O$)
RSBI – Rapid shallow breathing index (Rf/Vt) (breaths/l)
RSBIWA – Weight adjusted RSBI (breaths/l/kg)
MV – minute volume (l/min)
VA – alveolar minute volume (l/min)
EE – energy expenditure (kcal/min)
$FIO_2Set$ – Ventilator set inspired oxygen fraction.
VtSet – Ventilator set tidal volume (l)
PCSet – Ventilator set pressure control level ($cmH_2O$)
RfSet – Ventilator set respiratory frequency.
IESet – Ventilator set Inspiratory:Expiratory ratio
PEEPset – Ventilator set positive end expiratory pressure.

In addition to values recorded on a minute by minute basis, values from arterial blood gas input into the BC were recorded, these included $SaO_2$, $PaO_2$, $PaCO_2$ pH, BE, Hb, FMetHb, and FCOHb. Cardiac output values were either entered into the system, or estimated from IBW.

Ventilator mode – This was saved for both the mode reported by the ventilator, and that interpreted by the BC depending on the patient's breathing pattern.

In addition, pressure, flow, oxygenation and carbon dioxide waveforms were measured from the same recording sensors with a frequency of 50 Hz.

The following recordings were taken from the BC to analyse use of the system advice as described in section S.4 of this document. These variables describe the state of the system at any time point, and if advice presents the values of ventilator setting for this advice. The variables recorded are:

System status variables

- Advice provided, awaiting response.
- Patient not in steady state.
- ABG request provided, awaiting response.
- Typing in ABG.
- Performing ALPE.
- System paused, physiotherapy etc.
- Unsupported ventilator mode.
- Waiting, poor signal.
- Systems alarm status.

Advice variables

- $FIO_2$ advice
- Rf advice
- Pressure advice
- Vt advice
- PEEP advice
- Advice bounds exceeded
- Change mode advice

**S.4) Evaluation of advice use.**

Recording the time periods and nature of advice provided by the Beacon Caresystem along with the changes in ventilator settings allowed evaluation of how often advice was followed. To do so required definition of the different types of advice, along with definition of following or not following an advice. These definitions are provided here with the subsequent evaluation provided in table 5 of the main paper.

An advice was evaluated as "actionable" if it suggested changing ventilator settings from current conditions. The Beacon Caresystem provides advice to remain at current conditions and these were not defined as "actionable". In addition, the system can provide advice on a subset of ventilator settings, for example $FIO_2$ alone, with other settings suggested to remain as is. In this case only the specific setting is considered actionable. The function of the BC means that the patient is evaluated every minute. Advice calculated to be the same from minute to minute is not presented to the user every minute, and therefore counts as a single advice.

For "actionable" advice the following possibilities were defined

*Followed*: An advice is considered followed if acted upon within a 2 hour window of being presented. Advice is considered followed if the setting is adjusted in the same direction as the advice. If the advice includes changing two settings, for example both tidal volume and respiratory rate, then the advice is only considered followed if both are changed in the correct direction within this duration.

*Not acted on*: An advice is considered not acted on if the 2 hour window is exceeded without any ventilator change.

*Settings different than advised*: this situation is considered true if ventilator settings are changed in a different direction to those advised within the 2 hour window. If advice includes changing two settings, and only one is changed within the time window this is classified as Settings different than advised. The logic of this decision is that if advice suggests decreasing tidal volume but increasing respiratory rate, then only decreasing tidal volume is not a similar clinical intervention. The same applies for a combination of $FIO_2$ and PEEP advice.

*Remain*: Occasionally an advice to change settings is changed to an advice to remain at current settings during the 2hr window, thus becoming non-actionable. In this case the advice is classified as Remain and not evaluated as part of actionable advice.

*Cancelled*: If following advice a ventilator clinical event – extubation or mode change- occurs within the 2 hour window then the advice is considered no longer actionable and not evaluated as part of actionable advice.

Four types of advice were considered

1) Oxygen – This is advice in any ventilator mode on $FIO_2$, PEEP or a combination of these settings.
2) Pressure control – In pressure control mode advice can be for pressure level, respiratory frequency or a combination of these two settings.
3) Volume control – In volume control mode advice can be for tidal volume, respiratory frequency, or a combination of these two settings.
4) Pressure support – This is defined as advice on PS alone.

**S.5) References**


1. Patel, B., Mumby, S., Johnson, N. et al. Decision support system to evaluate ventilation in the acute respiratory distress syndrome (DeVENT study)—trial protocol. Trials 23, 47 (2022). https://doi.org/10.1186/s13063-021-05967-2.
2. Karbing DS, Spadaro S, Dey N, et al. An open-loop, physiologic model-based decision support system can provide appropriate ventilator settings. Crit Care Med 2018;46:e642–8.
3. Spadaro S, Karbing DS, Dalla Corte F, et al. An open-loop, physiological model based decision support system can reduce pressure support while acting to preserve respiratory muscle function. Journal of Critical Care, 2018, 48: 407–413.
4. Rees SE, Karbing DS. Determining the appropriate model complexity for patient-specific advice on mechanical ventilation. Biomed Tech 2017;62:183–98.
5. Rees SE, Kjærgaard S, Thorgaard P, Malczynski J, Toft E, Andreassen S. The Automatic Lung Parameter Estimator (ALPE) system: Non-invasive estimation of pulmonary gas exchange parameters in 10-15 minutes. Journal of Clinical Monitoring and Computing, 2002, Vol 17, No.1, pp 43-52.
6. Thomsen LP, Karbing DS, Smith BW, Murley D, Weinreich UM, Kjærgaard S, Toft E, Thorgaard P, Andreassen S, Rees SE. J Clin Monit Comput. 2013, June 27(3), 341-50.
7. Karbing DS, Kjærgaard S, Andreassen S, Espersen K, Rees SE. Minimal model quantification of pulmonary gas exchange in intensive care patients. Medical Engineering and Physics, 2011, 33_240-248.
8. Rees SE, Allerød C, Murley D, Zhao Y, Smith BW, Kjærgaard S, Thorgaard P, Andreassen S. Using physiological models and decision theory for selecting appropriate ventilator settings. Journal of Clinical Monitoring and Computing, 2006; Dec;20(6):421-429.
9. Rees SE, Spadaro S, Dalla Corte F, Dey N, Brohus JB, Scaramuzzo G, Lodahl D, Winding R, Volta CA, Karbing DS. Transparent decision support for mechanical ventilation using visualization of clinical preferences. Biomed Eng Online. 2022, in press
10. Gattinoni L, Tonetti T, Cressoni M, Cadringher P, Herrmann P, Moerer O, Protti A, Gotti M, Chiurazzi C, Carlesso E, Chiumello D, Quintel M. Ventilator-related causes of lung injury: the mechanical power Intensive Care Med. 2016 Oct;42(10):1567-1575.
11. Hess DR. Respiratory Mechanics in Mechanically Ventilated. Respiratory Care, 2014 59:1773-1794.
12. Pletsch-Assuncao R, Pereira MC, Ferreira JG, et al. Accuracy of invasive and noninvasive parameters for diagnosing ventilatory overassistance during pressure support ventilation. Crit Care Med. 2018;3:411–7.

**S.6) Acknowledgements**

**Main study contributors:**
**Dr Brijesh Patel**, Division of Anaesthetics, Pain Medicine, and Intensive Care, Department of Surgery and Cancer, Imperial College, London, UK
**Dr Sharon Mumby**, Airway Disease, National, Heart & Lung Institute, Imperial College, London, UK
**Mr Nicholas Johnson**, Imperial Clinical Trials Unit, Stadium House, 68 Wood Lane, London, W12 7RH
**Dr Emanuela Falaschetti**, Imperial Clinical Trials Unit, Stadium House, 68 Wood Lane, London, W12 7RH
**Dr Rhodri Handslip,** Division of Anaesthetics, Pain Medicine, and Intensive Care, Department of Surgery and Cancer, Imperial College, London, UK
**Dr Sunil Patel,** Division of Anaesthetics, Pain Medicine, and Intensive Care, Department of Surgery and Cancer, Imperial College, London, UK
**Dr Teresa Lee,** Division of Anaesthetics, Pain Medicine, and Intensive Care, Department of Surgery and Cancer, Imperial College, London, UK
**Dr Martin S Andersen,** Respiratory and Critical Care Group, Department of Health Science and Technology, Aalborg University, Aalborg, Denmark
**Professor Ian Adcock**, Airway Disease, National, Heart & Lung Institute, Imperial College, London, UK.
**Professor Danny F McAuley**, Wellcome-Wolfson Institute for Experimental Medicine, Queen's University, Belfast, UK
**Professor Masao Takata**, Division of Anaesthetics, Pain Medicine, and Intensive Care, Department of Surgery and Cancer, Imperial College, London, UK
**Dr Thomas Staudinger,** Medical University of Vienna, Department of Medicine I, Waehringer Guertel 18-20, A-1090 Vienna, Austria
**Dr Dan S.Karbing**, Respiratory and Critical Care Group, Department of Health Science and Technology, Aalborg University, Aalborg, Denmark
**Professor Matthieu Jabaudon**, Department of Perioperative Medicine, University Hospital of Clermont-Ferrand, GReD, Université Clermont Auvergne, CNRS, INSERM, Clermont-Ferrand, France
**Dr Peter Schellongowski**, Medical University of Vienna, Department of Medicine I, Waehringer Guertel 18-20, A-1090 Vienna, Austria
**Professor Stephen E Rees**, Respiratory and Critical Care Group, Department of Health Science and Technology, Aalborg University, Aalborg, Denmark

***DeVENT study contributors: Imperial College London** – Mayur Murali. **Imperial Clinical Trials Unit** – Ana Boshoff; Mary Cross. **Royal Brompton & Harefield Hospitals** – Mahitha Gummaddi; Vicky Thwaites; Natalie Dormand; Anelise Catelan Zborowski ; Geraldine Sloane; Mathew Varghese; Kamal Liniyage; Adam Pitt; Daniele Cristiano; the clinical and research staff at Royal Brompton Hospital critical care. **Medical University Vienna** – Thomas Staudinger; Elisabeth Lobmeyr; Nina Buchtele, Oliver Robak, Andja Bojic, Alexander Hermann, Valerie Weinberger, Barbara Krieger, Bernhard Nagler; Julia Cserna; the clinical and research staff at MUV. **Clermond-Ferrand University Hospital** – Benjamin Rieu, Jean-Baptiste Joffredo, Thomas Costille, Emmanuel Futier, Julien Amat; Dominique Morand; Raiko Blondonnet; Jules Audard; The clinical and research staff at CHU Clermont-Ferrand and Universite Clermont Auvergne. **Mermaid Care A/S** – Soren Thomsen; Jakob B Brohus. **Independent Data Monitoring and Ethics Committee** – Professor Charlotte Summers (Chairperson); Dr Steve Harris; Professor David Harrison (independent statistician); Professor Mark Griffiths. **Independent Trial Steering Committee** – Professor Giacomo Bellani (Chairperson); Professor Dan Brodie; Professor John Laffey; Mrs Yosien Burke (patient and public representative)

**Funding**
The European Commission Horizon 2020 Fast Track to Innovation Programme will be providing the research costs for the DeVENT study (Reference EU project 853850 - ECMO-BIOMARKER). The funder had no role in

the trial design, collection, management, analysis or interpretation of data, writing of reports or submission for publication.
